# Prevalence of Long COVID-associated symptoms in adults with and without SARS-CoV-2 infection in Germany: Results of the population-based study “Corona Monitoring Nationwide 2021/22” (RKI-SOEP-2)

**DOI:** 10.1101/2023.09.12.23295426

**Authors:** Christina Poethko-Müller, Ana Ordonez-Cruickshank, Julia Nübel, Giselle Sarganas, Antje Gößwald, Lorenz Schmid, Angelika Schaffrath Rosario, Jens Hoebel, Martin Schlaud, Christa Scheidt-Nave

## Abstract

**Background:** Controlled population-based studies on long-term health sequelae of SARS-CoV-2 can help to identify clinical signs specific to “Long COVID” and to evaluate this emerging public health challenge.

**Aim:** To examine prevalence differences of Long COVID-associated symptoms among adults with and without SARS-CoV-2 infection in Germany.

**Methods:** This population-based, retrospective study (11/2021-2/2022) included 7,683 working aged adults (18-65 years), a subset of the Corona Monitoring Nationwide study in Germany. Prior SARS-CoV-2 infection was defined based on self-reported PCR-confirmed infections and IgG-antibody dried blood spot testing. Participants answered a questionnaire including 19 common symptoms of Long COVID experienced in the six months preceding the survey. We estimated population-weighted prevalence of (1) individual symptoms, and (2) ≥1 symptom, with and without impact on work ability, by infection status within strata of sex, age group, income and comorbidity. We calculated model-adjusted prevalence differences and the probability that symptoms among infected are attributable to infection.

**Results:** 12 of 19 symptoms showed a significantly higher prevalence in infected than non-infected participants, including fatigue (27.5% versus 18.3%; p<0.001), concentration problems (22.2% vs. 13.1%; p<0.001), shortness of breath (15.6% vs. 7.5%; p<0.001), and smell and taste disorder (10% vs. 1.2%; p<0.001). ≥1 symptom with impact on work ability was more prevalent following infection (16.0% vs. 12.2%; p=0.06) with a model-adjusted prevalence difference of 3.8% (95%-CI -0.5-8.0).

**Conclusion:** We observed a rather small excess prevalence attributable to SARS-CoV-2 infection. However, the absolute number of persons places great demands on the health care system and may affect economic productivity.

## Introduction

Globally, more than 769 million cases of Severe Acute Respiratory Syndrome Coronavirus 2 (SARS-CoV-2) infection and almost 7 million deaths have been reported until August 2023. In Germany there were over 38 million cases of confirmed SARS-CoV-2 infection as of August 2023 (1). SARS-CoV-2 has challenged healthcare systems, not only because of the impact of the acute disease but also because of its long-term health sequelae known as ‘post COVID-19 condition’. The World Health Organization (WHO) defines post COVID-19 as a condition usually occurring three months after a confirmed or probable SARS-CoV-2 infection, with symptoms that may be new onset, fluctuate or persist from the initial illness generally impacting everyday functioning, lasting for at least two months, and not explainable by alternative diagnoses (2). ‘Long COVID’ is a term initially coined by patients in the social media (3). According to the current state of knowledge, it can be assumed that Long COVID is a multifactorial post-acute infection syndrome with complex underlying disease mechanisms (4, 5).

Large population-based epidemiological studies have identified several commonly reported symptoms of Long COVID. These include fatigue, shortness of breath, memory, and concentration problems (2, 6-9). Nevertheless, it has been extremely challenging to provide estimates of prevalence, determinants and public health impact of Long COVID in epidemiological studies or meta-analyses from primary studies. Besides limited insight into the disease mechanisms and course of disease, this has been hampered by highly variable study methodology, in particular lack of standardization of outcome and exposure measurements, data source, study population and of missing control groups (9, 10). As many symptoms of Long COVID are prevalent in the general population irrespective of a SARS-CoV-2 infection, lack of an adequate control group is likely to inflate prevalence estimates of symptoms (7, 11). Few studies have been population-based and also included a contemporary control group which would permit calculation of differences between infected and non-infected persons adjusting for covariables, i.e. the prevalence of symptoms that can be attributed to COVID-19 (12).

The present analysis used nationwide data from a study embedded in a long-standing population-based household panel in Germany to assess the period prevalence of Long COVID-associated symptoms among adults of working age according to SARS-CoV-2 infection status at the turn of 2021/2022. We aimed to estimate the period prevalence of symptoms attributable to SARS-CoV-2 infection and the absolute number of adults affected by symptoms.

## Methods

### Study design and study population

Data was derived from the second wave of the cross-sectional population-based “Corona Monitoring nationwide” (RKI-SOEP-2) study, a cooperative project of the Robert Koch Institute (RKI), the Socio-Economic Panel (SOEP) at the German Institute for Economic Research (DIW Berlin), the Institute for Employment Research (IAB), and the Research Center of the Federal Office for Migration and Refugees (BAMF-FZ). The study was embedded in the SOEP panel, a German nationwide dynamic cohort based on random population samples, and comprised a sample of all individuals aged 14 years and older who participated in the 2021 SOEP-2 wave. Participation was mainly in November and December 2021 – a period with predominance of the Delta variant in Germany. Details of the study design have been previously published (13).

The present analysis is confined to 7,683 adults of working age (18-65 years) who participated in SOEP-2, had complete questionnaire and laboratory information on SARS-CoV-2 infection status and Long COVID-associated symptoms, no self-reported PCR-confirmed SARS-CoV-2 infection within four weeks preceding the survey, and no history of previous hospitalization because of SARS-CoV-2 infection (see Flow chart, Supplement Figure 1).

### Measures and definitions

Participants provided a self-collected dried blood sample, which were tested for immunoglobulin G (IgG) antibodies against the spike protein and the nucleocapsid protein of SARS-CoV-2 (Euroimmun QuantiVac IgG-ELISA; cut-off ≥ 11 and Anti-SARS-CoV-2-NCP; cut-off adapted for dried blood spot testing ≥ 0.95) (14).

Information on former infection, hospitalization due to COVID-19, Long COVID-associated symptoms, functional limitation (work ability), and vaccination status were collected by a self-administered questionnaire (written form or online). Information on chronic non-communicable diseases (NCDs) had previously been self-reported in SOEP questionnaires (data from the 2019 SOEP wave or the latest available data from earlier waves) and income data from 2021.

Definition of exposure, i.e. SARS-CoV-2 infection status, was based on self-reported history of a PCR-confirmed infection and antibody test results. History of ‘known infection’ was assumed when a positive PCR-test result dating ≥ 4 weeks before study participation was reported; infection status of participants with self-report of never having had a SARS-CoV-2 infection before was defined as ‘unnoticed infection’ if they were tested seropositive for anti-N IgG antibodies (or positive for anti-S1 IgG antibodies if unvaccinated); otherwise, they were categorized as ‘non-infected’.

The presence of Long COVID-associated symptoms was assessed based on a list of 19 common symptoms of Long COVID, selected based on available evidence from previous epidemiological studies on Long COVID (complete list of symptoms in Figure 1).

**Figure 1.**
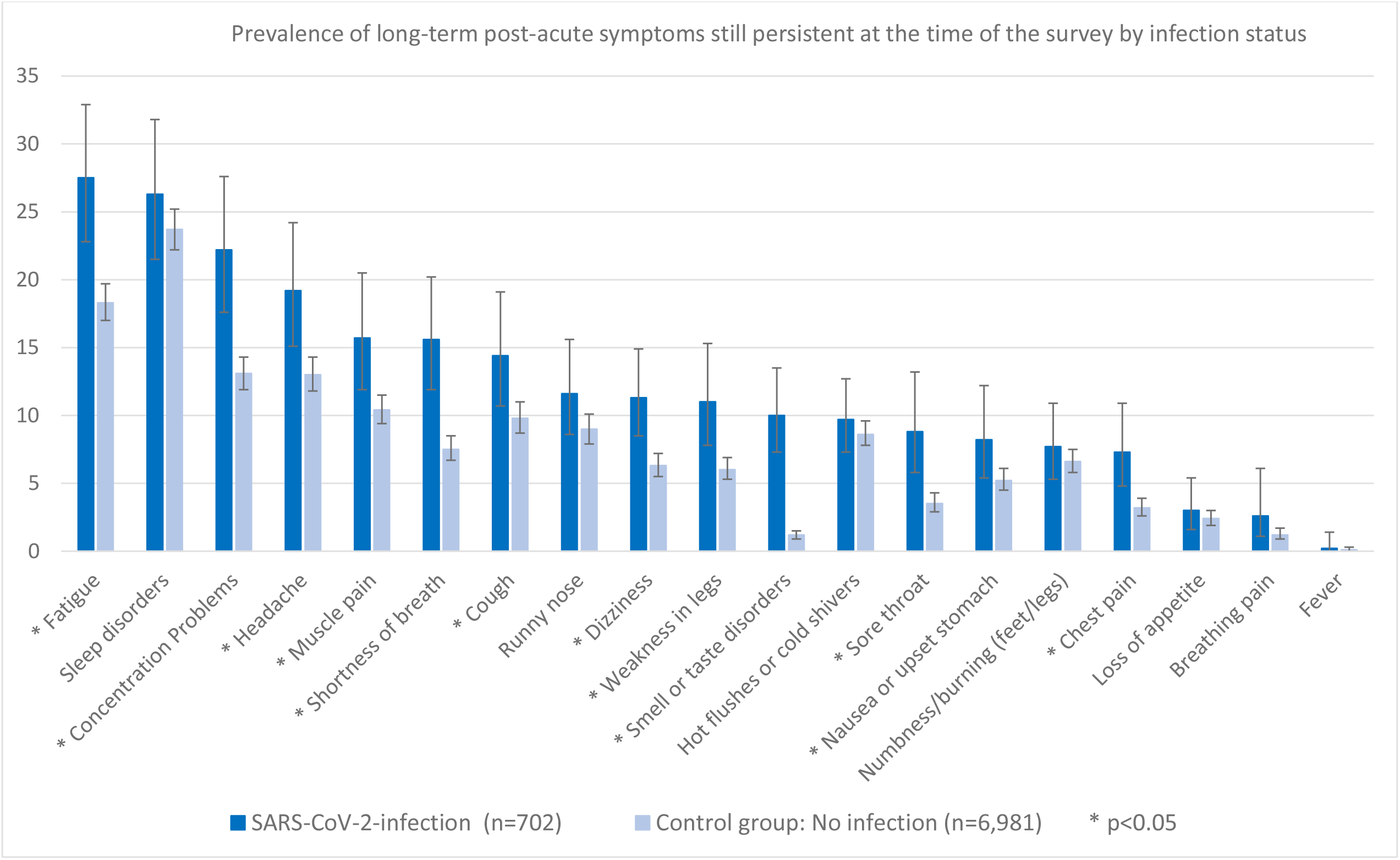

Long COVID-associated symptoms were defined if participants reported to have experienced ≥1 permanent or recurrent symptom in the past 6 months which still persisted at the time of the survey.

Having Long COVID-associated symptoms with impact on work ability was defined if participants reported the presence of ≥1symptom impacting everyday functioning. The question was “Have these complaints affected the following areas?”. The response categories were tailored to adults of working age: work; job search; vocational training/further education; attending school/studying.

Self-reported history of NCDs included physician-diagnosed diabetes, asthma, heart disease, cancer, stroke, migraine, high blood pressure, depression, dementia, and joint disease. The total number of diagnoses was categorized into 0 or ≥1.

Income was calculated by dividing the household–s total disposable income by the square root of the number of household members and categorized into low (quintile 1), middle (quintiles 2-4), or high (quintile 5) (15).

### Statistical analysis

Analyses were performed using Stata survey procedures (version 16.1, StataCorp, College Station, TX, USA) to account for household clustering and weighting. Survey-specific weighting factors were used to compensate for sampling design and non-random non-response. The weighting strategy considered different selection processes (contactability and participation) from the gross sample to the net sample at both the household and the individual level and adjusted to known population distributions with regard to socio-demographic structure (age and gender distribution, socio-demographic status, residential area, and migration history) (13).

We compared the prevalence of Long COVID-associated symptoms between persons with (known or unnoticed) and without SARS-CoV-2 infection. Analyses were stratified by sex, age group, presence of NCDs and income. Results are presented in unweighted numbers (n) and weighted percentages (%) with 95% logit confidence intervals (95%-CI). We calculated weighted prevalence differences between infected and non-infected persons (in percentage points), which are unadjusted estimates of the risk that an infected person develops Long COVID, i.e. Long COVID-associated symptoms that are indeed attributable to the infection. Model-adjusted prevalence differences were based on a logistic regression with symptoms as target variable and infection status, age group, sex, NCDs and income as independent variables, including interaction terms of infection status with the other covariates. Prevalence differences and CIs were calculated using the Stata ‘margins’ and ‘regpar’ modules (16). An analysis using a linear binomial model yielded similar results (data not shown).

We estimated the model-adjusted probability that symptoms among infected persons are indeed Long COVID, i.e. attributable to the infection (attributable risk among exposed, ARE) with 95%-CI using the ‘punaf’ module (16), which is equivalent to dividing the model-adjusted prevalence difference by the prevalence among infected.

The absolute number of Long COVID in the German population of working age was estimated by multiplying the prevalence difference (i.e. the risk of Long COVID in infected) with the prevalence of previous SARS-CoV-2 infection.

## Results

### Baseline characteristics

Overall, women were slightly overrepresented compared to men (Table 1). Nearly half of the participants were 50-65 years of age (45.8%), about one third were 34-49 years of age, and 22% were 18-33 years of age. Mean age of the study population was 45.3 years. Half of the participants reported one or more NCDs (50.0%). The proportion of participants with known SARS-CoV-2 infection was 6.1%, plus 2.5% with unnoticed infection. The median time since acute infection in participants with known infection was 294 days (range 29-689 days). Most participants were vaccinated against COVID-19 at least once (93.4%) (Table 1).

**Table 1.** Baseline characteristics of the study population 18 to 65 years of life in Germany 2021/2022, overall (column %) and population-weighted prevalence of SARS-CoV-2 infection status.

|  |  |  |  |  |  |  |  |
| --- | --- | --- | --- | --- | --- | --- | --- |
|  |  | Infection status |  |  |  |  |  |
|  | Total n, unweighted (column %, weighted) | Known infection > 4 weeks ago |  | Unnoticed infection |  | No infection |  |
|  |  | n (unweighted) | % weighted [95%CI] or mean | n (unweighted) | % weighted [95%CI] or mean | n (unweighted) | % weighted [95%CI] or mean |
| **Total (age 18-65 yol)** | 7,683 | 535 | 6.1 [5.2-7.0] | 167 | 2.5 [2.0-3.2] | 6,981 | 91.4 [90.3-92.4] |
| **Sex** |  |  |  |  |  |  |  |
| Male | 3,415 (44.4) | 231 | 6.2 [5.1-7.5] | 75 | 3.0 [2.1-4.2] | 3,109 | 90.8 [89.1-92.2] |
| Female | 4,268 (55.6) | 304 | 5.9 [5.0-7.0] | 92 | 2.0 [1.5-2.6] | 3,872 | 92.1 [90.9-93.2] |
| **Age** |  |  |  |  |  |  |  |
| 18-33 | 1,698 (22.1) | 137 | 6.4 [5.0-8.3] | 45 | 3.4 [2.3-5.0] | 1,516 | 90.2 [87.9-92.1] |
| 34-49 | 2,463 (32.1) | 194 | 7.5 [6.0-9.5] | 53 | 2.8 [1.9-4.0] | 2,216 | 89.7 [87.5-91.5] |
| 50-65 | 3,522 (45.8) | 204 | 4.6 [3.6-5.7] | 69 | 1.6 [1.1-2.4] | 3,249 | 93.8 [92.5-95.0] |
| Age in years (mean) | 45.3 (weighted 43.0) | 43.1 | 40.8 | 44.4 | 39.9 | 45.4 | 43.2 |
| **Number of NCDs** |  |  |  |  |  |  |  |
| 0 | 3,676 (50.0) | 261 | 6.0 [4.9-7.3] | 91 | 2.9 [2.1-4.0] | 3,324 | 91.0 [89.4-92.4] |
| ≥1 | 3,680 (50.0) | 237 | 5.7 [4.6-6.9] | 70 | 1.9 [1.3-2.7] | 3,373 | 92.4 [91.0-93.6] |
| **Income** |  |  |  |  |  |  |  |
| Low | 1,022 (14.3) | 79 | 6.8 [4.7-9.8] | 37 | 3.8 [2.3-6.0] | 906 | 89.4 [86.0-92.1] |
| Middle | 4,142 (58.0) | 295 | 6.3 [5.2-7.7] | 78 | 2.3 [1.7-3.2] | 3,769 | 91.3 [89.8-92.7] |
| High | 1,983 (27.7) | 121 | 4.3 [3.2-5.8] | 32 | 1.7 [0.9-3.2] | 1,830 | 94.0 [92.1-95.4] |
| **Time since infection** |  |  |  |  |  |  |  |
| Median (days) | Range 29-689 days | Unweighted 294 | Weighted 286 |  |  |  |  |
| **Vaccination status** |  |  |  |  |  |  |  |
| Not vaccinated | 504 (6.6) | 101 | 15.7 [11.1-21.8] | 40 | 10.5 [6.4-16.9] | 363 | 73.7 [66.4-79.9] |
| vaccination ≥1 dose | 7,178 (93.4) | 433 | 5.3 [4.5-6.3] | 127 | 1.9 [1.5-2.5] | 6,618 | 92.7 [91.7-93.7] |

Numbers do not add up to total due to missing values in single variables (available-case analysis): Number of missings NCD: n=327; Income n=536

### Pattern of individual long COVID-associated symptoms

12 of 19 preselected symptoms were significantly more prevalent in the group with previous SARS-CoV-2 infection compared to the non-infected group. In descending order of prevalence (Figure 1, Table 1 SUPPLEMENT), these symptoms included fatigue (27.5% versus 18.3%; p<0.001), difficulties in concentration (22.2% vs. 13.1%; p<0.001), headaches (19.2% vs. 13.0%; p=0.0031), muscle pain (15.7% vs. 10.4%; p=0.0053), shortness of breath (15.6% vs. 7.5%; p<0.001), cough (14,4% vs. 9,8%; p=0.0176), dizziness (11.0% vs. 6.3%; p=0.0002), weakness in legs (11.0% vs. 6.0%; p=0.0012), smell or taste disorder (10.0% vs. 1.2%; p<0.001). No significant differences were observed for sleep disorders, for numbness or burning sensations in feet or legs, loss of appetite, pain when breathing or signs of acute infection such as runny nose, hot flushes or cold shivers, and fever (Figure 1).

### Prevalence of any Long COVID-associated symptom

The overall population-weighted prevalence of ≥1 Long COVID-associated symptom among adults with SARS-CoV-2 infection was 56.4% (95%-CI 50.7-62.0) (Table 2); the respective proportion of symptoms in non-infected persons was significantly lower (47.7%; 95%-CI 46.0-49.5%), prevalence difference 8.7%; p=0.0044). The prevalence of ≥1 symptoms was higher in infected and non-infected females (67.6% and 56.3) in comparison to men (46.9% and 39.1%) and was only statistically different among females (prevalence difference 11.3%; p=0.0041). Prevalence was higher in 50- to 65-year-olds than in younger adults and differed between infected and non-infected in this age group (prevalence difference 18.2%, p<0.001). Prevalence was higher in infected and non-infected persons with comorbidity. However, the prevalence difference was higher in those without comorbidity.

**Table 2.** Population-weighted prevalence of one or more persistent long COVID-like symptoms, by infection status and sociodemographic and health-related determinants in adults 18-65 years of life (yol) in Germany 2021/22.

|  | Stratified by infection status |  |  |  |  |  |  |  |  |  |  |
| --- | --- | --- | --- | --- | --- | --- | --- | --- | --- | --- | --- |
| Baseline characteristics | Known infection > 4 weeks ago (n=535) |  | Unnoticed infection (n=167) |  | Total infected (n=702) |  | Not infected (n=6,981) |  | Prevalence difference (total infected versus not infected) | p-value*<br>* total infected versus not infected | Model-adjusted***<br>prevalence difference |
|  | n** | Prevalence in % (95%-CI) | n*<br>* | Prevalence in % (95%-CI) | n** | Prevalence in % (95%-CI) | n** | Prevalence in % (95%-CI) | Difference in percentage points % (95%-CI) |  | Difference in percentage points % (95%-CI) |
| **Total 18-65 yol n=7,683** | 334 | 60.9 [54.3-67.0] | 80 | 45.6 [34.3-57.4] | 414 | 56.4 [50.7-62.0] | 3,394 | 47.7 [46.0-49.5] | 8.7 [2.7-14.5] | 0.0044 | 12.1 [6.6-17.5] |
| **Sex** |  |  |  |  |  |  |  |  |  |  |  |
| Male | 119 | 51.0 [41.1-60.9] | 31 | 38.6 [23.4-56.4] | 150 | 46.9 [38.4-55.7] | 1,177 | 39.1 [36.6-41.7] | 7.8 [-1.3-16.8] | 0.0846 | 10.1 [1.0-19.1] |
| Female | 215 | 71.4 [63.1-78.5] | 49 | 56.3 [42.6-69.1] | 264 | 67.6 [60.4-74.1] | 2,217 | 56.3 [54.0-58.7] | 11.3 [3.9-18.5] | 0.0041 | 14.2 [6.7-21.5] |
| **Age** |  |  |  |  |  |  |  |  |  |  |  |
| 18-33 | 73 | 52.8 [40.1-65.1] | 18 | 46.9 [27.9-66.9] | 91 | 50.8 [39.9-61.5] | 682 | 43.5 [40.1-47.1] | 7.2 [-4.4-18.6] | 0.2170 | 9.2 [-2.5-20.6] |
| 34-49 | 117 | 60.6 [49.6-70.6] | 18 | 24.1 [12.9-40.4] | 135 | 50.8 [41.4-60.1] | 993 | 45.3 [42.2-48.5] | 5.4 [-4.4-15.2] | 0.2751 | 1.9 [-7.2-11.1] |
| 50-65 | 144 | 69.9 [58.-,79.3] | 44 | 73.6 [57.5-85.2] | 188 | 70.9 [61.5-78.7] | 1,719 | 52.7 [50.1-55.3] | 18.2 [9.0-27.1] | <0.001 | 22.6 [12.7-32.1] |
| **Number of NCDs** |  |  |  |  |  |  |  |  |  |  |  |
| 0 | 147 | 57.3 [47.6-66.5] | 39 | 39.4 [25.3-55.5] | 186 | 51.4 [43.1-59.7] | 1,259 | 36.8 [34.3-39.3] | 14.7 [5.9-23.2] | <0.001 | 20.1 [12.1-27.8] |
| >=1 | 166 | 68.5 [59.0-76.7] | 38 | 58.2 [40.9-73.7] | 204 | 65.9 [57.6-73.3] | 2,015 | 60.4 [58.0-62.8] | 5.5 [-2.6-13.6] | 0.1973 | 3.2 [-4.8-11.2] |
| **Income** |  |  |  |  |  |  |  |  |  |  |  |
| Low | 37 | 44.8 [28.1-62.9] | 18 | 62.1 [39.2-80.7] | 55 | 51.0 [36.6-65.2] | 508 | 53.9 [49.0-58.7] | -2.9 [-18.1-12.4] | 0.7087 | 7.9 [-6.5-21.9] |
| Middle | 199 | 68.1 [60.0-75.3] | 36 | 42.5 [27.5-59.1] | 235 | 61.3 [53.8-68.3] | 1,843 | 47.7 [45.3-50.0] | 13.6 [6.0-21.1] | <0.001 | 17.1[9.7-24.2] |
| High | 70 | 49.2 [38.4-60.1] | 17 | 32.4 [13.4-59.7] | 87 | 44.4 [33.5-55.9] | 803 | 41.9 [38.3-45.6] | 2.5 [-9.4-14.3] | 0.6776 | 1.8 [-8.4-11.9] |

\*\*n = Number of participants with symptoms; \*\*\*Model variables: Infection status plus sex, age group, presence of NCDs, income and their interaction with infection; Number of missings for NCD: n=327; Income n=536

Prevalence difference was significant in the middle-income group, without differences in the low- and high-income groups. Compared to non-infected adults, the prevalence was consistently higher in those with infection; the difference was highest in 50- to 65-year-olds (18.2%). The overall model-adjusted prevalence difference was 12.1% (95%-CI 6.6-17.5) (Table 2).

### Prevalence of any Long COVID-associated symptom with functional impact

The overall population-weighted prevalence of ≥1 Long COVID-associated symptom with functional limitation in the ability to work or study was 16.0% (95%-CI 12.2-20.7) in those infected and 12.2% (95%-CI 11.0-13.5%) in non-infected persons (Table 3), with a prevalence difference of 3.8 percent points (95%-CI -0.6-8.2; p=0.06), equal to the model-adjusted prevalence difference (95%-CI -0.5-8.0). The unadjusted prevalence difference was equal in men and women (4.0%), whereas the model-adjusted difference was somewhat higher in women (4.6%; 95%-CI -1.3-10.4 vs. 2.9%; 95%-CI -2.9-8.8). The unadjusted prevalence difference was highest in 34- to 49-year-olds (5.9%; 95%-CI -1.5-13.3), while the adjusted difference was highest in the two highest age groups (both 4.4%).

**Table 3.** Population-weighted prevalence of one or more persistent long COVID-like symptom with functional limitation, by infection status and sociodemographic and health-related determinants in adults 18-65 years of life (yol) in Germany 2021/22.

| Stratified by infection status |  |  |  |  |  |  |  |  |  |  |  |
| --- | --- | --- | --- | --- | --- | --- | --- | --- | --- | --- | --- |
| Baseline characteristics | Known infection > 4 weeks ago (n=535) |  | Unnoticed infection (n=167) |  | Total infected (n=702) |  | Not infected (n=6,981) |  | Prevalence difference (total infected versus not infected) | p-value*<br>* total infected versus not infected | Model-adjusted*** prevalence difference |
|  | n** | Proportion in % or mean (95%-CI) | n** | Proportion in % or mean (95%-CI) | n** | Proportion in % (95%-CI) | n** | Proportion in % (95%-CI) | Difference in percentage points % (95%-CI) |  | Difference in percentage points % (95%-CI) |
| **Total 18-65 yol n= 7,124** | 85 | 16.4 [11.9,22.0] | 25 | 15.3 [8.9,24.8] | 110 | 16.0 [12.2,20.7] | 733 | 12.2 [11.0,13.5] | 3.8 [-0.6-8.2] | 0.0600 | 3.8 [-0.5-8.0] |
| **Sex** |  |  |  |  |  |  |  |  |  |  |  |
| Male | 33 | 16.4 [10.2,25.4] | 9 | 9.8 [4.1,21.8] | 42 | 14.3 [9.4,21.2] | 272 | 10.3 [8.7,12.2] | 4.0 [-2.1-10.0] | 0.1548 | 2.9 [-2.9-8.8] |
| Female | 52 | 16.3 [11.1,23.3] | 16 | 23.3 [12.4,39.3] | 68 | 18.1 [13.0,24.6] | 461 | 14.1 [12.4,16.0] | 4.0 [-2.0-10.0] | 0.1611 | 4.6 [-1.3-10.4] |
| **Age** |  |  |  |  |  |  |  |  |  |  |  |
| 18-33 | 18 | 13.3 [6.6,24.9] | 9 | 21.7 [9.7,41.6] | 27 | 16.2 [9.7,25.9] | 244 | 14.6 [12.3,17.2] | 1.7 [-6.8-10.0] | 0.6884 | 2.1 [-7.7-11.7] |
| 34-49 | 36 | 19.6 [12.1,30.1] | 8 | 9.4 [3.8,21.4] | 44 | 17.0 [11.0,25.2] | 194 | 11.1 [9.0,13.5] | 5.9 [-1.5-13.3] | 0.0754 | 4.4 [-3.0-11.7] |
| 50-65 | 31 | 15.2 [9.4,23.7] | 8 | 12.6 [4.8,29.4] | 39 | 14.5 [9.5,21.6] | 295 | 11.3 [9.7,13.1] | 3.3 [-3.0-9.5] | 0.2656 | 4.4 [-2.3-11.1] |
| **Number of NCDs** |  |  |  |  |  |  |  |  |  |  |  |
| 0 | 35 | 14.2 [8.4,23.0] | 12 | 8.7 [3.6,19.7] | 47 | 12.4 [7.9,19.1] | 239 | 8.2 [6.8,9.9] | 4.2 [-1.5-9.9] | 0.0918 | 3.1 [-2.0-8.2] |
| >=1 | 47 | 21.2 [14.4,30.1] | 10 | 24.0 [11.8,42.7] | 57 | 22.0 [15.7,29.8] | 459 | 16.6 [14.7,18.7] | 5.4 [-2.0-12.6] | 0.1161 | 4.3 [-3.1-11.7] |
| **Income** |  |  |  |  |  |  |  |  |  |  |  |
| Low | 12 | 22.2 [9.8,42.9] | 5 | 16.3 [5.6,39.1] | 17 | 20.1 [10.5,35.0] | 179 | 21.2 [17.4,25.5] | -1.1 [-13.9-11.7] | 0.8694 | 4.2 [-9.5-17.8] |
| Middle | 54 | 16.3 [11.2,23.1] | 11 | 14.5 [6.6,29.0] | 65 | 15.8 [11.3,21.7] | 401 | 11.6 [10.1,13.3] | 4.2 [-1.2-9.5] | 0.0948 | 4.3 [-0.9-9.6] |
| High | 13 | 8.5 [4.4,15.8] | 5 | 16.0 [4.0,46.6] | 18 | 10.6 [5.4,19.8] | 105 | 7.1 [5.2,9.7] | 3.5 [-3.7-10.7] | 0.2683 | 1.9 [-4.0-7.7] |

\*\*n = Number of participants with symptoms; \*\*\*Model variables: Infection status plus sex, age group, presence of NCDs, income and their interaction with infection; Number of missings for NCD: n=309 ; Income n=495

Prevalence difference by comorbidity was similar in those without and with comorbidity (4.2 and 5.4 percent points). The pattern of a social gradient with a higher prevalence in lower income groups was apparent in both infected and non-infected, although the 95% CI overlapped in the infected group (Table 3).

### Long COVID-associated symptoms attributable to SARS-CoV-2 infection and estimated number of persons affected in Germany

The model-adjusted probability that Long COVID-associated symptoms in the infected were attributable to the infection was 20.2% (95%-CI 12.4-27.3) (Table 4), and the corresponding probability for symptoms with functional limitation was 23.5% (95%-CI -0.9-41.9). The estimated risk of Long COVID in the population was 0.7% for Long COVID-associated symptoms without and 0.3% for symptoms with functional limitation (Table 4), corresponding to an estimated number of 390.000 of 18- to 65-year-olds with Long COVID, and of 170.000 with Long COVID impacting work ability.

**Table 4.** Attributable risk of Long COVID among exposed and risk of Long COVID in the German population 2021/22.

|  |  |  |
| --- | --- | --- |
|  | One or more persistent Long COVID-associated symptoms | One or more persistent Long COVID-associated symptoms, with functional limitation |
| **Attributable risk of Long COVID among exposed**<br>(Probability for an infected person with Long COVID-associated symptoms that the symptoms are indeed Long COVID, i.e. have been caused by the infection*,**) | 20.2% (95%-CI 12.4-27.3) | 23.5% (95%-CI -0.9-41.9) |
| **Estimated risk of Long COVID in the German population (infected and not infected) aged 18 to 65 yol\*** | Descriptive: 0.7%<br>Model-adj**: 1.0% | Descriptive: 0.3%<br>Model-adj**: 0.3% |
| **Estimated number of cases\*, \*\*\*** | ~ 390,000 | ~ 170,000 |
| *At the turn of 2021/2022 (Delta-to-Omicron transition)<br>**Model variables: Infection status plus sex, age group, presence of NCDs, income and their interaction with infection status<br>***Estimated risk (descriptive) multiplied by population size 18-65 yol (as of 31.12.2020: 52,2 million) |  |  |

## Discussion

### Key results

This nationwide cross-sectional study of adults aged 18-65 years in Germany was conducted at the turn of 2021/2022 to assess the prevalence of Long COVID-associated symptoms based on comparative analysis of persons with previous SARS-CoV-2 infection and non-infected controls.

Among 19 preselected symptoms previously shown to be frequently present in the post-acute phase of COVID-19 (≥ 4 weeks after infection), 12 symptoms were significantly more prevalent among persons with than without previous SARS-CoV-2 infection, with the most considerable prevalence differences observed for smell and taste disorders, fatigue, concentration problems, and shortness of breath. The prevalence of ≥1 symptom was consistently higher in persons with than without SARS-CoV-2 infection. The overall population-weighted prevalence difference, which is the estimated risk of Long COVID (defined as ≥1 symptom) in the infected, was 8.7%. The model-adjusted prevalence difference was 12.1%. The prevalence difference of ≥ 1 symptoms impacting on working or studying abilities was smaller (3.8%). According to these data, about 390,000 of 18- to 65-year-old persons in Germany had persistent health complaints at the turn of 2021/2022 that can be attributed to SARS-CoV-2 infection, and in about 170,000 persons, these health complaints impaired working or studying abilities.

### Comparison with other studies

#### Pattern of individual symptoms

Many national and international studies support the very high symptom burden of fatigue, difficulties in concentration and shortness of breath, and smell and taste disorder after SARS-CoV-2 infection (6, 9, 17-20).

A systematic review and meta-analysis on persistent symptoms up to 12 months after infection calculated the prevalence of dyspnea on exertion (34%; 95%-CI 0.20-0.94); difficulties in concentration (32%; 0.16-0.52), fatigue (31%; 0.22-0.40) and arthromyalgia (28%; 0.09-0.6) (21). Estimates in this meta-analysis are higher than in this study, possibly due to the exclusion of individuals >65 yol here. The wide range of prevalence estimates across studies may result from differences between countries and the severity of COVID-19 infection in study populations, but most probably also indicate the use of non-harmonized measurement tools and the lack of comparison against control groups (22).

An early regional study in Germany identified anosmia, ageusia, fatigue, and shortness of breath as the most common, persisting symptoms at months 4 and 7 among patients with confirmed SARS-CoV-2 infection in 2020 (23). Another uncontrolled regional study in southwest Germany estimated the prevalence of post-acute symptoms and symptom clusters of 18- to 65-year-old adults 6-12 months after confirmed SARS-CoV-2 infection. Adjusting for self-reported symptoms before infection, this previous study observed an overall prevalence of ≥1 post-acute symptoms of 28.5% and a minimum estimate of 6.5% when additionally considering potential non-response bias (24). The symptom clusters fatigue and neurocognitive impairment were frequently reported by about one third of the study participants and contributed most to reduced health recovery and working capacity (24). Although the prevalence estimates from this large regional German study and our nationwide study cannot be directly compared, both studies consistently highlight the importance of fatigue and neurocognitive impairment as possible persistent symptoms following SARS-CoV-2 infection in the working-age population.

A population-based multi-center cohort study of adults 18 years and older in Germany examined health consequences among individuals about 1 to 1.5 years after SARS-CoV-2 infection, compared with non-infected adults (25). Consistent with the results of the present study, this previous report found a higher prevalence of a number of individual symptoms among persons with than without previous SARS-CoV-2 infection (28). The observed prevalence difference for the presence of ≥ 1 out of 18 preselected recurrently occurring or persisting symptoms was 10.7 percent points and thereby comparable to the prevalence difference calculated in our nationwide study.

#### Prevalence of Long COVID-associated symptoms

Our prevalence estimates of 56% for at least one persistent symptom after SARS-CoV-2 infection is somewhat higher than the results of an extensive systematic review and meta-analysis (including studies up to January 2022), with a mean prevalence of 45% for at least one prolonged symptom after about four months (21). However, the comparison of results is hindered by heterogeneous study designs, differences in follow-up durations, and measurement methods. In addition, overall symptom burden and subjective awareness of health complaints may vary by study population (e.g. depending on demographic characteristics such as age and sex).

In a longitudinal study from the Netherlands, 41% of participants had at least one new onset somatic symptom (out of a list of 23 symptoms) of moderate severity at 90–150 days of follow-up after documented SARS-CoV-2 infection, compared with 29% of non-infected controls (matched for age and sex) resulting in a prevalence difference of 11% in 2020 and 2021 (6). These findings are comparable to the adjusted prevalence difference observed in the present study from Germany, which was 12% at the end of 2021.

Notably, prevalence estimates are highly dependent on the definition and the background prevalence of long-term symptoms in the non-infected control population, and a lack of universal agreement on the clinical definition of long COVID, variation in data sources, and methodology produce discordant results (26). A majority of studies focused only on the presence of specific symptoms – without considering daily functioning or health-related quality of life. Considering the importance of impairment in daily life, we applied different definitions to assess the different degrees of severity. The adjusted prevalence difference for ≥ 1 long-lasting symptom was 12.1 percent points. However, the proportion of persons who reported impairment of professional and educational daily tasks due to the experienced symptoms was 16.0% versus 12.2% with a difference of 3.8%. A recent nationwide prospective cohort study from Norway based on health record data in primary care observed a small excess in symptoms typical of Long COVID among persons with previously documented SARS-CoV-2 infection compared to non-infected controls amounting to 0.05-2.5% depending on the type of symptom and the length of follow-up (9). Taken together, results from recent controlled studies in various countries support findings of the present analysis indicating a relatively small prevalence difference in persons with persisting post-acute symptoms impairing work ability. Nevertheless, given the high number of persons infected with SARS-CoV-2, the absolute individual and social burden to healthcare systems and to the employment market will be high as demonstrated by the estimated numbers of 390.000 affected adults of working age in Germany at the turn of 2021/2022 who had persisting post-acute symptoms attributable to non-severe SARS-CoV-2 infection, and about 170.000 of these with impairment of working or studying ability.

### Strengths and limitations

#### Strengths

The present study was based on a household panel representative of the German population. Notably, the study included a non-infected control group. The combination of information derived from questionnaires and serological analyses allowed us to limit possible misclassification of persons with unnoticed SARS-CoV-2 infections as not infected. The database permitted the estimation of symptom prevalence according to exposure status and the calculation of prevalence differences and attributable risks.

#### Limitations

The main limitation of the present study is its observational and cross-sectional design precluding causal inferences and giving rise to potential selection and recall bias.

Some participants were unaware of their infection (‘unnoticed infection’) and could still have been in the acute phase of their infection. The NICE guideline for Long COVID definition, which requires an elapsed period of four weeks after acute infection, would not be fulfilled for these participants (27). However, since participants with unnoticed infection were identified based on IgG N-antibodies (28) known to occur after some delay in acute infection, this group is less likely to include individuals with acute infection. Although there might be residual misclassification of individuals with acute SARS-CoV-2-infection, the amount of any overestimation should be minor because unnoticed infections are likely to have been asymptomatic or mild and less likely to lead to Long COVID (29). An underestimation of the prevalence of Long COVID-associated symptoms may have occurred due to selective non-response of less healthy individuals, the exclusion of institutionalized subjects and cases hospitalized because of COVID-19.

Our study was not designed to analyze possible effects of vaccination on Long COVID. We obtained self-reported information on vaccination status, but it was difficult to use this information in multivariable analyses for several reasons. The number of vaccine doses received and the timing in relation to infection was highly variable resulting in a large number of subgroups of small sample size. In addition, no date of infection was known for persons with unnoticed SARS-CoV-2-infection. Most of all, as vaccine prioritization strategies were necessary up to the second half of 2021, vaccination status correlated with a higher risk of Long COVID-associated symptoms due to higher comorbidity and higher age (30). Additional analyses including at least one dose of COVID-19-vaccine in our models showed no relevant change of main results.

## Conclusion

Not all Long COVID-associated symptoms in persons with previous SARS-CoV-2 infection are due to the infection. We observed a rather small excess prevalence of symptoms attributable to SARS-CoV-2 infection, in particular when additionally considering the impact of symptoms on working or studying abilities. However, the absolute number of persons affected not only causes great individual suffering but also places great demands on the health care system and may affect economic productivity.

## Ethical issues

Approval was obtained from the ethics committee of the Berlin State Chamber of Physicians (reference ID Eth-33/20 as of September 21, 2021). All participants provided informed consent.

We declare that the planning conduct and reporting of studies was in line with the Declaration of Helsinki, as revised in 2013.

The RKI-SOEP-2 study was funded by the German Federal Ministry of Health (project number ZMI1-2521COR305). The funders had no role in the design and conduct of the study, in the collection, management, analysis and interpretation of the data, in the preparation, review or approval of the manuscript, or in the decision to submit the manuscript for publication.

## Data Availability

The data cannot be made publicly available because informed consent from participants did not cover the public deposition of data. However, the data underlying the analysis in this article is archived in the SOEP Research Data Centre (https://www.diw.de/en/diw_01.c.601584.en/data_access.html) in Berlin and can be accessed on site upon reasonable request.

## Acknowledgements

We would like to thank all our colleagues at the Robert Koch Institute (RKI), the Socio-Economic Panel (SOEP) at the German Institute for Economic Research (DIW Berlin), the Institute for Employment Research (IAB) and the Research Centre of the Federal Office for Migration and Refugees (BAMF-FZ) for their support and cooperation. Special thanks go to the staff of DIW Econ GmbH for carrying out the weighting. We also thank the employees of the infas Institute for Applied Social Sciences who contributed to the planning and implementation of fieldwork and data collection. We sincerely thank all study participants for their willingness to participate.

---

**Fig. S1.**
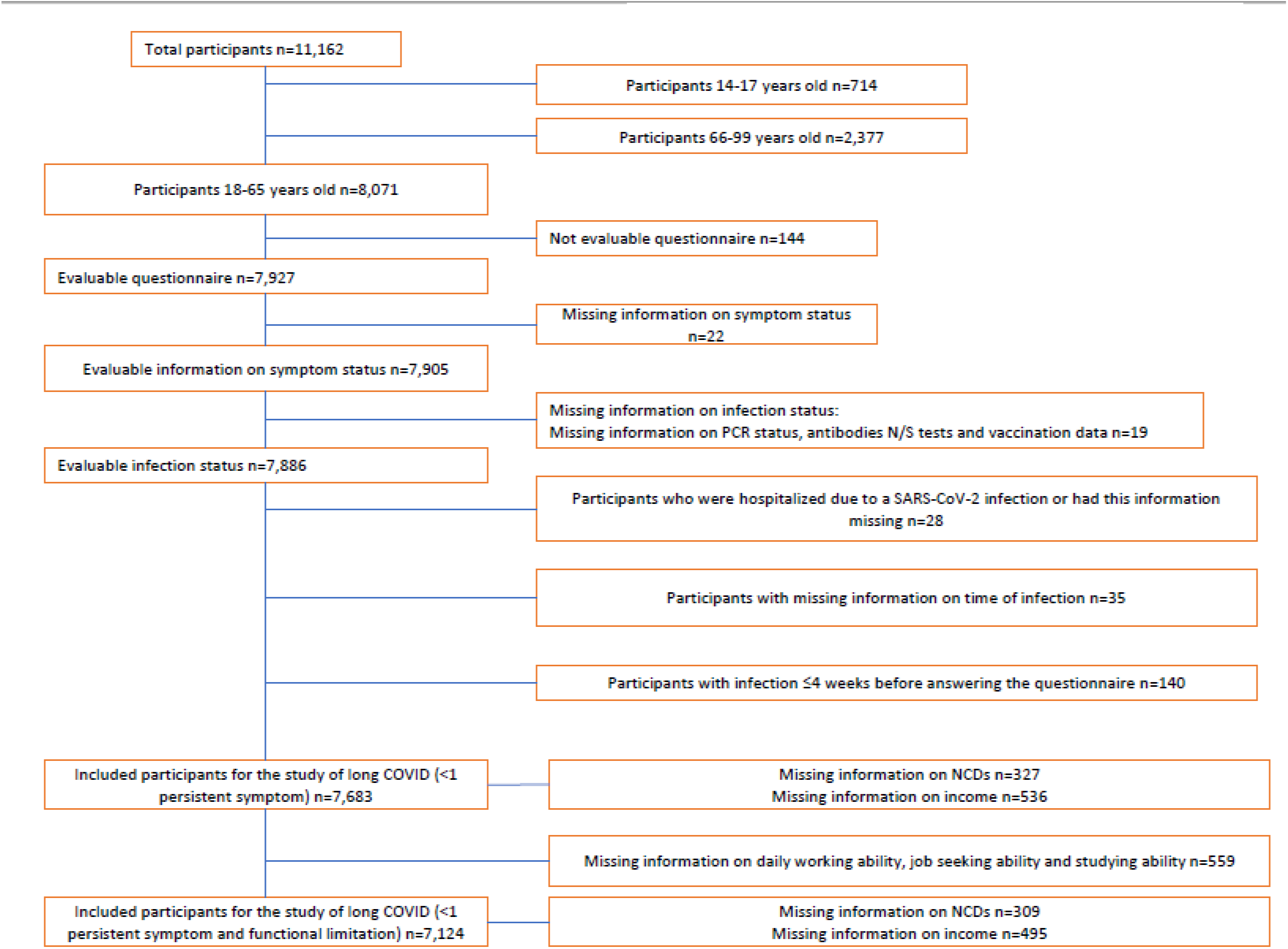
Flowchart: Numbers of exclusions.

**Table S1.** Prevalence of long-term post-acute symptoms persisting up to present (known and unnoticed infection versus no infection)

|  |  |  |  |  |  |  |  |  |
| --- | --- | --- | --- | --- | --- | --- | --- | --- |
| Symptoms | Overall<br>n<br>(total<br>n=7.683) | SARS-CoV-2-infection<br>≥4<br>weeks<br>ago [%]<br>(n=702) | 95%-CI |  | Control<br>group:<br>No<br>infection<br>(n=6,981) | 95%-CI |  | p-Value:<br>Infected vs.<br>non-infected<br>people |
| Fatigue | 1.481 | 27,5 | 22,8 - | 32,9 | 18,3 | 17,0 - | 19,7 | <0.001 |
| Sleep disorders | 1.912 | 26,3 | 21,5 - | 31,8 | 23,7 | 22,2 - | 25,2 | 0,3107 |
| Concentration Problems | 1.048 | 22,2 | 17,6 - | 27,6 | 13,1 | 11,9 - | 14,3 | <0.001 |
| Headache | 997 | 19,2 | 15,1 - | 24,2 | 13 | 11,8 - | 14,3 | 0,0031 |
| Muscle pain | 859 | 15,7 | 11,9 - | 20,5 | 10,4 | 9,4 - | 11,5 | 0,0053 |
| Shortness of breath | 625 | 15,6 | 11,9 - | 20,2 | 7,5 | 6,7 - | 8,5 | <0.001 |
| Cough | 678 | 14,4 | 10,7 - | 19,1 | 9,8 | 8,7 - | 11,0 | 0,0176 |
| Runny nose | 662 | 11,6 | 8,6 - | 15,6 | 9 | 7,9 - | 10,1 | 0. 1150 |
| Dizziness | 539 | 11,3 | 8,5 - | 14,9 | 6,3 | 5,5 - | 7,2 | 0,0002 |
| Weakness in legs | 511 | 11 | 7,8 - | 15,3 | 6 | 5,3 - | 6,9 | 0,0012 |
| Smell or taste disorders | 164 | 10 | 7,3 - | 13,5 | 1,2 | 0,9 - | 1,5 | <0.001 |
| Hot flushes or cold shivers | 783 | 9,7 | 7,3 - | 12,7 | 8,6 | 7,8 - | 9,6 | 0,4325 |
| Sore throat | 269 | 8,8 | 5,8 - | 13,2 | 3,5 | 2,9 - | 4,3 | <0.001 |
| Nausea or upset stomach | 405 | 8,2 | 5,4 - | 12,2 | 5,2 | 4,5 - | 6,1 | 0,0435 |
| Numbness or burning sensation in feet or legs | 519 | 7,7 | 5,3 - | 10,9 | 6,6 | 5,8 - | 7,5 | 0,4354 |
| Chest pain | 259 | 7,3 | 4,8 - | 10,9 | 3,2 | 2,6 - | 3,9 | <0.001 |
| Loss of appetite | 160 | 3 | 1,6 - | 5,4 | 2,4 | 1,9 - | 3,0 | 0,5257 |
| Breathing pain | 97 | 2,6 | 1,1 - | 6,1 | 1,2 | 0,9 - | 1,7 | 0,1208 |
| Fever | 5 | 0,2 | 0,001 - | 1,4 | 0,1 | 0,0 - | 0,3 | 0,4213 |

